# Ethnic differences in *SERPINA1* allele frequencies may partially explain national differences in COVID-19 fatality rates

**DOI:** 10.1101/2020.08.24.20179226

**Authors:** Guy Shapira, Noam Shomron, David Gurwitz

**Affiliations:** Sackler Faculty of Medicine, Tel Aviv University, Tel Aviv 69978, Israel; Sagol School of Neuroscience, Tel Aviv University, Tel Aviv 69978, Israel; Edmond J Safra Center for Bioinformatics, Tel Aviv University, Tel Aviv 69978, Israel

**Keywords:** alpha-1 antitrypsin, SERPINA1, ethnic variation, COVID-19, rs28929474, rs17580

## Abstract

COVID-19 infection and fatality rates vary considerably between countries. We present preliminary evidence that these variations may in part reflect ethnic differences in the frequencies of polymorphic alleles of *SERPINA1*, coding for alpha-1 antitrypsin, the major blood serine protease inhibitor.

## Introduction

Differences between countries in state-mandated border closures, lockdowns, and social distancing regulations provide obvious reasons for the great variation between countries in COVID-19 infection rates and morbidity. Differences in cultural customs have also been attributed a role. For example, wearing face masks was common in some Asian countries prior to the current pandemic, and this may have contributed to lower COVID-19 infection and fatality rates in these countries [1]. Genetic variations between ethnic groups have also been proposed as a factor that contributes to differences in COVID-19 epidemiology. The relatively low rates of COVID-19 infection and mortality in East and Southeast Asian countries, including Japan, China, South Korea, Thailand, Vietnam, Cambodia, and Malaysia, are notable, and remain little understood [2]. Relatively low COVID-19 infection and mortality rates are also notable for several Sub-Saharan African countries. While these rates may be due to lower testing for SARS-CoV-2 and fewer international arrivals, they may also reflect genetic variations between the populations. However, at the time of this writing, no studies have reported associations of genetic variations between populations with COVID-19 infection and fatality rates.

The serine protease TMPRSS2 is well-established as essential for priming SARS-CoV-2 cell entry, following binding to its ACE2 cell membrane receptor [3]. Accordingly, the synthetic serine protease inhibitor nafamostat mesylate and camostat mesylate were shown to inhibit TMPRSS2 [4] and are undergoing clinical trials in COVID-19 patients. The major human blood serine protease inhibitor (serpin) is alpha-1 antitrypsin, which is encoded by the *SERPINA1* gene and produced in the liver, similar to many blood proteins. Comparing regions in Italy, differential alpha-1 antitrypsin genetic deficiency rates were proposed to correlate with COVID-19 infection. This was based on higher rates of both alpha-1 antitrypsin deficiency (carriers of *SERPINA1* rs28929474, termed PiZ) and COVID-19 infection rates in Lombardy, Italy than in central and southern Italian regions [5]. We therefore examined a possible association between the distributions of common *SERPINA1* single nucleotide polymorphisms (SNPs) underlying alpha-1 antitrypsin deficiency and COVID-19 epidemiology on a global scale.

## Methods

National allele frequency estimates for the alpha-1 antitrypsin PiS *(SERPINA1* rs17580) and PiZ *(SERPINA1* rs28929474) alleles were obtained from Blanco et al. [6]. National parameters were obtained from the United Nations database and data.worldbank.org. For combining the effects of the PiS and PiZ alleles on alpha-1 antitrypsin serum concentration, their frequencies were summed, weighted by their approximate effect size (50% and 90% respectively, according to Mitchell et al. [7]). Confounder correction and assessment of individual factor effect sizes were done using one-way ANCOVA (Analysis of Covariance). All statistical analyses were made in the R programming environment. Country-specific epidemiological parameters of the ongoing COVID-19 pandemic were compared between all countries using data from the Johns Hopkins University Coronavirus Resource Center [8] (accessed August 23, 2020). We focused on countries with a population of over one million in order to reduce noise, e.g. from small island nations.

## Results and discussion

We examined if population variations in human serine protease inhibitor (serpin) genes may help explain the lower COVID-19 infection rates and fatalities in East and Southeast Asia and Sub-Saharan Africa, compared to other regions. Among the 43 human serpin genes, *SERPINA1*, coding for alpha-1 antitrypsin, the major blood serine protease inhibitor, showed the most striking variations in SNP frequencies between Europeans and non-Europeans. We examined in particular variations in *SERPINA1* rs28929474 (the PiZ allele). This allele is recognized as the major allele responsible for antitrypsin deficiency in humans, and was recently suggested to account for the higher COVID-19 infection rates in Lombardy compared with other Italian regions [5]. *SERPINA1* rs28929474 was about 8-fold less frequent in East and Southeast Asian than in South European populations (17 compared to 2 alleles per 1,000 individuals). For the milder deficiency allele, *SERPINA1* rs17580 (PiS allele), the disparity was even greater (86 compared to 5 alleles per 1,000 individuals, over a 17-fold reduction). The combined analysis of the global PiZ and PiS allele frequencies is shown in **Figure 1a**.

**Figure 1.**
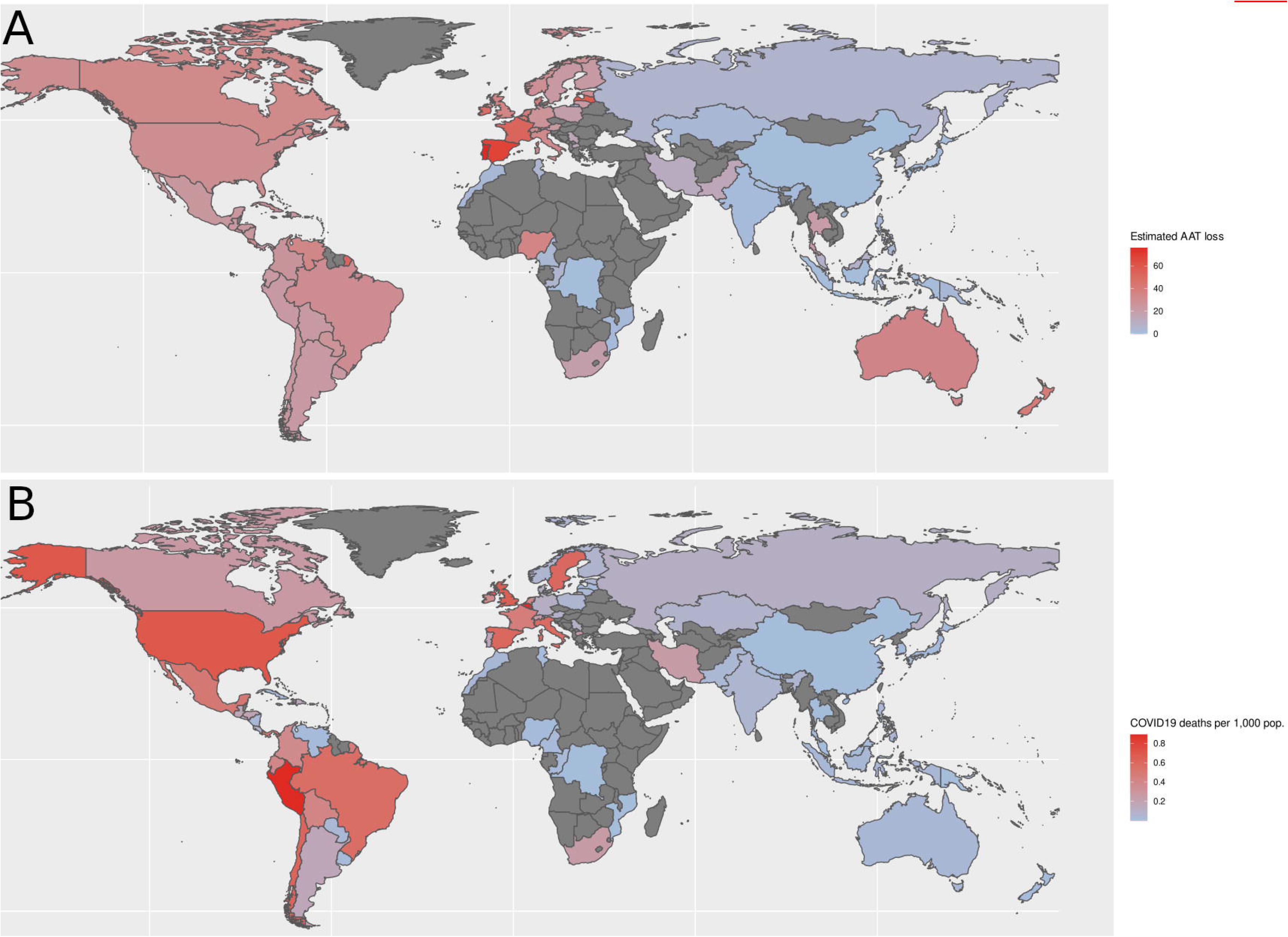
Demographics of national alpha-1 antitrypsin deficiency allele frequencies and COVID-19 fatality rates per 1,000 population. (**a**) Estimated population frequencies of alpha-1 antitrypsin deficiency alleles PiZ and PiZ (*SERPINA1* rs28929474 and rs17580, respectively). Aggregated estimates were taken from Blanco et al. [6], and summed proportionally to their respective effect sizes on alpha-1 antitrypsin serum concentration. (**b**) COVID-19 fatality rates per 1,000 population were accessed from the Johns Hopkins University Coronavirus Resource Center [8] (August 21, 2020).

Our comparison of epidemiological data showed that nearly all the countries reporting >100 COVID-19 mortalities per million population were in Europe or the Americas. In contrast, East and Southeast Asian, and also Sub-Saharan African countries were the predominant countries with low mortality rates per million population (**Figure 1b; Table S1**). The COVID-19 population-adjusted fatality rates for Thailand, Vietnam, and Cambodia were 0.8, 0.2, and <0.1 per million, respectively [6]; low COVID-19 fatality rates were also notable for several Sub-Saharan African nations, such as Congo, and Mozambique (2 and 0.7 per million, respectively). Thus, the difference is up to ~100-fold compared with some European and American countries. The far lower population-adjusted burden of COVID-19 in many Asian and Sub-Saharan African countries remains notable when comparing total infections per one million people (**Table S1**).

It could be argued that the lower COVID-19 infection and fatality rates in many Asian and Sub-Saharan countries reflect their lower human development index (HDI), urbanization level, and volume of international travel, as well as lower proportions of elderly people. However, our analysis indicates that this is not the case) (the analysis is available by request from the authors). Moreover, infection and fatality rates remain among the highest globally in several South-American countries with low HDI and relatively low proportions of elderly persons. The striking difference in the COVID-19 burden between South American countries and between East and Southeast Asian countries therefore seems to be best explained by ethnic genetic variation. Until additional candidate alleles are found to better explain such national differences, the population variations in the allele frequencies of alpha-1 antitrypsin deficiency present a possible genetic explanation for the varying national COVID-19 burden.

We applied estimated country-specific frequencies of alpha-1 antitrypsin deficiency [6] to analyze COVID-19 fatalities per thousand inhabitants vs. estimated alpha-1 antitrypsin deficiency in 67 countries (**Fig. 2**). The Pearson correlation of R=0.56 (P=8.7e-7) suggests an association of alpha-1 antitrypsin deficiency with COVID-19 fatalities. To eliminate the confounding effects of such factors as urbanization and age distribution, we used a one-way ANCOVA (Analysis Of Covariance) that accounts for HDI. The results showed a significant association after correction (P=0.0002; the full analysis is available in the supplementary methods file). Considering the above, we propose that the low carrier frequency of alpha-1 antitrypsin deficiency PiZ and PiS alleles in East and Southeast Asian and Sub-Saharan African countries may help explain the lower COVID-19 infection and mortality rates in these regions.

**Figure 2.**
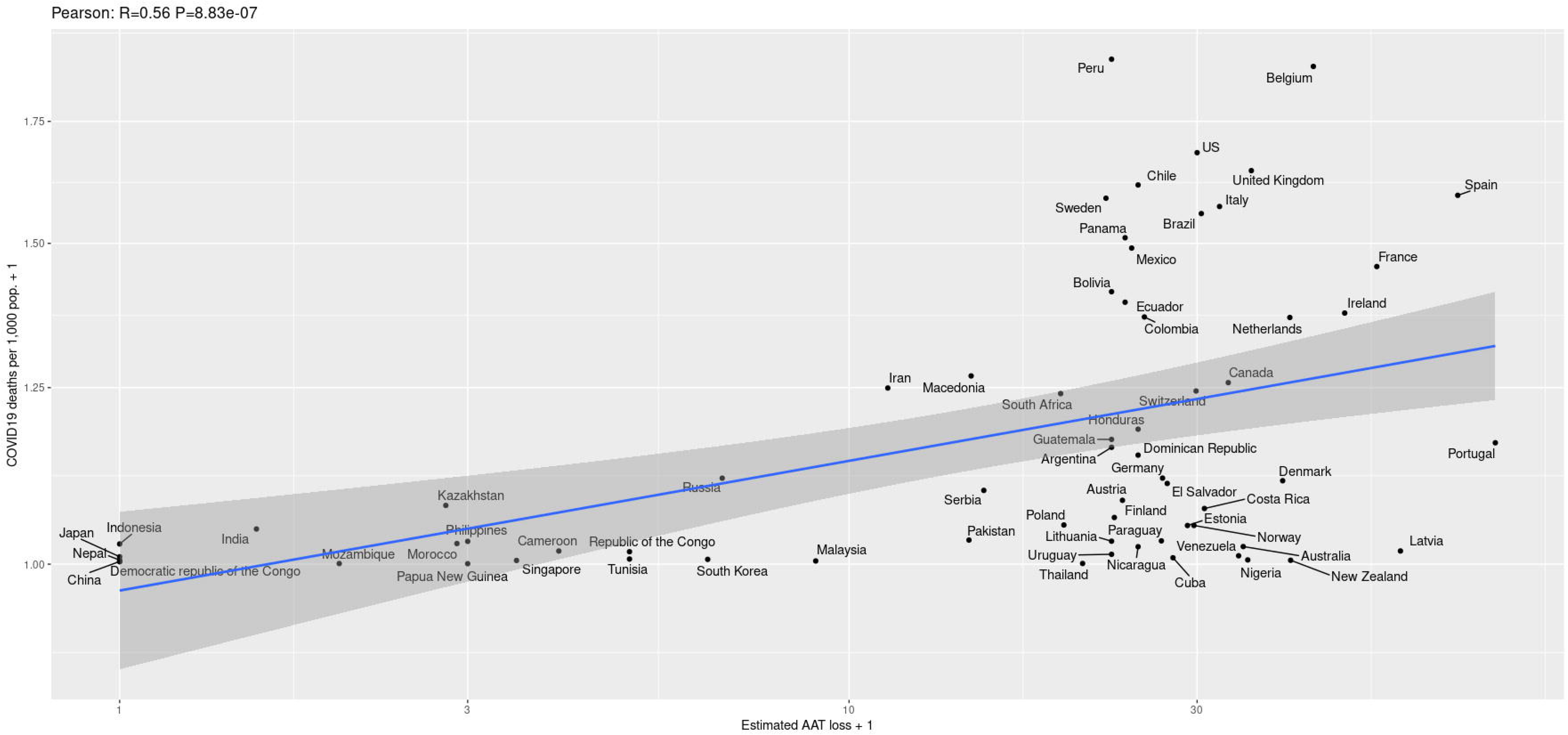
Positive correlations between estimated national alpha-1 antitrypsin deficiency allele frequencies and COVID-19 fatality rates per 1,000 population. The Pearson correlation of R=0.56 (P=6.48e-7) suggests an association of national rates of alpha-1 antitrypsin deficiency with the national rates of COVID-19 fatalities. See Figure 1 for details on data sources. Only countries with at least a population of >1 million are included.

In addition to being the major blood serpin, alpha-1 antitrypsin is known as a potent anti-inflammatory protein and a key regulator of the human acute phase immune response. McElvaney et al. [9] recently reported elevated plasma alpha-1 antitrypsin levels in COVID-19 patients. This suggests the involvement of alpha-1 antitrypsin in the acute phase immune response to SARS-CoV-2 infection, which might be insufficient for protection against COVID-19.

The bacillus Calmette-Guérin (BCG) vaccination was proposed as protective from severe COVID-19 [10], while it was shown to increase alpha-1 antitrypsin blood levels in BCG vaccinated individuals together with trained immunity [11]. Notably, a recent study (posted as preprint and not yet published at submission of this manuscript) demonstrated in vitro inhibition of human TMPRSS2 by alpha-1 antitrypsin, thus suggesting that it might be effective as a COVID-19 treatment [12]. Another recent preprint showed that alpha-1 antitrypsin may inhibit in vitro SARS-CoV-2 infection in human airway epithelium at physiological concentrations [13]. A clinical trial of inhaled alpha-1 antitrypsin in COVID-19 patients is currently recruiting participants (NCT04385836). If alpha-1 antitrypsin deficiency will be established as a key genetic risk factor for severe COVID-19, population-wide screening for this deficiency will be desirable, for identification and prioritizing for vaccination, once a vaccine becomes available.

## Data Availability

All data and statistical tests can be found in: https://gitlab.com/shep/serpina1_covid_supplementary

## Conflict of interest

The authors declare that no conflicts of interest exist.

## Acknowledgements

DG is supported by the Yoran Institute for Human Genome Research at Tel Aviv University. The NS lab is partially supported by the Adelis Foundation. The authors thank Cindy Cohen for professional editorial assistance.

## Data availability

A complete reproducible analysis is available at: https://gitlab.com/shep/serpina1_covid_supplementary

## Supplementary Information

Table S1: COVID19 epidemiological data

Table S2: The dataset used for the analysis, including national alpha-1 antitrypsin allele frequencies and national indicators.

Supplementary Methods: See https://gitlab.com/shep/serpina1_covid_supplementary

## Notes

### Competing Interest Statement

The authors have declared no competing interest.

### Funding Statement

DG is supported by the Yoran Institute for Human Genome Research at Tel Aviv University. The team of NS is partially supported by the Adelis Foundation.

### Author Declarations

No approval required; no personal human data included.

## References

[1] Iwasaki A, Grubaugh ND. Why does Japan have so few cases of COVID-19? EMBO Mol Med. 2020; 12(5):e12481. doi: 10.15252/emmm.202012481.

[2] Gaye B. et al. Socio-demographic and epidemiological consideration of Africa’s COVID-19 response: what is the possible pandemic course? Nat Med. 2020; 26(7):996-999. doi: 10.1038/s41591-020-0960-y.

[3] Hoffmann M. et al, SARS-CoV-2 Cell Entry Depends on ACE2 and TMPRSS2 and Is Blocked by a Clinically Proven Protease Inhibitor. Cell. 2020; 181(2):271-280.e8. doi: 10.1016/j.cell.2020.02.052.

[4] Yamamoto M. et al. The Anticoagulant Nafamostat Potently Inhibits SARS-CoV-2 S Protein-Mediated Fusion in a Cell Fusion Assay System and Viral Infection In Vitro in a Cell-Type-Dependent Manner. Viruses. 2020; 12(6):629. doi: 10.3390/v12060629.

[5] Vianello A, Braccioni F. Geographical Overlap Between Alpha-1 Antitrypsin Deficiency and COVID-19 Infection in Italy: Casual or Causal? Arch Bronconeumol. 2020 May 31:S0300-2896(20)30169-1. doi: 10.1016/j.arbres.2020.05.015. Online ahead of print.

[6] Blanco I et al. Alpha-1 antitrypsin Pi*SZ genotype: estimated prevalence and number of SZ subjects worldwide. Int J Chron Obstruct Pulmon Dis. 2017; 12:1683-1694. doi: 10.2147/COPD.S137852.

[7] Mitchell EL, Khan Z. Liver Disease in Alpha-1 Antitrypsin Deficiency: Current Approaches and Future Directions [published correction appears in Curr Pathobiol Rep. 2018;6(1):97]. Curr Pathobiol. Rep. 2017;5(3):243-252. doi:10.1007/s40139-017-0147-5

[8] Dong E, Du H, Gardner L. An interactive web-based dashboard to track COVID-19 in real time. Lancet Infect Dis. 2020;20(5):533-534. doi: 10.1016/S1473-3099(20)30120-1.

[9] McElvaney O.J. et al. Characterization of the Inflammatory Response to Severe COVID-19 Illness. Am J Respir Crit Care Med. 2020 Jun 25. doi: 10.1164/rccm.202005-1583OC. Online ahead of print.

[10] Yitbarek K, Abraham G, Girma T, Tilahun T, Woldie M. The effect of Bacillus Calmette-Guérin (BCG) vaccination in preventing sever infectious respiratory diseases other than TB: Implications for the COVID-19 pandemic. Vaccine. 2020 Aug 10:S0264-410X(20)31049-5. doi: 10.1016/j.vaccine.2020.08.018. Online ahead of print.

[11] Cirovic B et al. BCG Vaccination in Humans Elicits Trained Immunity via the Hematopoietic Progenitor Compartment. Cell Host Microbe. 2020; 28(2):322-334.e5. doi: 10.1016/j.chom.2020.05.014.

[12] Nurit P. Azouz, Andrea M. Klingler, Marc E. Rothenberg. Alpha 1 Antitrypsin is an Inhibitor of the SARS-CoV2-Priming Protease TMPRSS2. bioRxiv 2020.05.04.077826; doi: https://doi.org/10.1101/2020.05.04.077826

[13] Wettstein L. et al. Alpha-1 antitrypsin inhibits SARS-CoV-2 infection. bioRxiv 2020.07.02.183764; doi: https://doi.org/10.1101/2020.07.02.183764

